# Posterior Globe Geometry in Vitreoretinal Disease

**DOI:** 10.64898/2026.02.01.26345318

**Authors:** João Heitor Marques, Ana Sofia Martins, Pedro Manuel Baptista, Bernardete Pessoa, João Melo Beirão

## Abstract

**Purpose:** To investigate whether the geometric shape of the globe, sagittal length (SL) and Posterior Radius of curvature (rPC), differ between patients with rhegmatogenous retinal detachment (RD) and macular hole (MH), and to determine if these parameters offer better diagnostic differentiation than axial length (AL) alone.

**Methods:** This retrospective study included 20 MH and 20 RD patients. Groups were axial-length matched. Ocular biometry was performed with ultrasound to measure AL and SL. The PR was calculated. Due to the non-normal distribution of the data, the Mann-Whitney U test was used for continuous variables and Fisher’s Exact Test for gender distribution.

**Results:** There was no significant difference between the MH and RD groups in terms of age (median: 33.8 vs 32.6 years; p=1.00), gender (p=0.341), or AL (27.65 mm vs 28.58 mm; p=0.39). However, RD eyes showed significantly higher SL (median: 25.70 mm vs 22.90 mm; p=0.001) and a significantly flatter rPC (median: 28.25 mm vs 24.30 mm; p < 0.001).

**Conclusion:** RD eyes exhibit a distinct vertical equatorial expansion and posterior flattening that is not present in MH eyes, despite similar axial lengths. These geometric differences suggest that SL and PR are structural risk factors in vitreoretinal disease.

## Introduction

In ophthalmology, axial length (AL) has long been the primary biometric marker for assessing the risk of retinal pathologies, particularly in myopic eyes [1]. However, the human eye is a three-dimensional structure that can expand asymmetrically. High axial myopia often involves the development of posterior staphylomas or irregular globe shapes that are not captured by a one-dimensional AL measurement [2]. Retinal Detachment (RD) and Macular Hole (MH) are both vision-threatening conditions caused by vitreoretinal traction, yet they occupy different anatomical sites (the periphery vs. the center). We hypothesize that the underlying geometry of the globe, specifically its sagittal length (SL) and the radius of curvature of the posterior segment (PR), differ significantly between these groups, even when their AL is comparable. This study aims to quantify these differences to better understand the structural predispositions for these pathologies.

## Methods

### Population and Design

A retrospective analysis was conducted in Centro Hospitalar Universitário de Santo António, a terciary reference center for vitreoretinal pathology. The study was approved by the Institutional Board and Ethics Committee with the number 2022.213 (171-DEFI/174-CE). We included 40 patients divided into two groups: 20 with MH and 20 with RD. Only one eye per patient was analyzed (the diseased eye or the first affected eye in case of bilateral disease).

### Biometric Measurement

All subjects underwent ocular ultrasound with Ellex Eye Cubed® and a 15Mhz probe, by a single experienced investigator that was blind to the patients’ group. Measurments are illustrated in Figure 1.AL was measured from the corneal vertex to the retinal surface along the main anatomical axis. A cross-sectional line was considered 22mm behind the corneal vertex and SL was measured in the vertical and horizontal axis, the average value of these two measurments was considered for analysis. To calculate the posterior curvature radius (rPC), three points of the limit of the vitreous cavity where considered: 1 and 2 where it crosses the 22mm line and 3 as the most posterior point. Three points define a unique circumference, and its radius (rPC) may be calculated using the width of the sides together with the area of a triangle defined by those 3 points.

**Figure 1.**
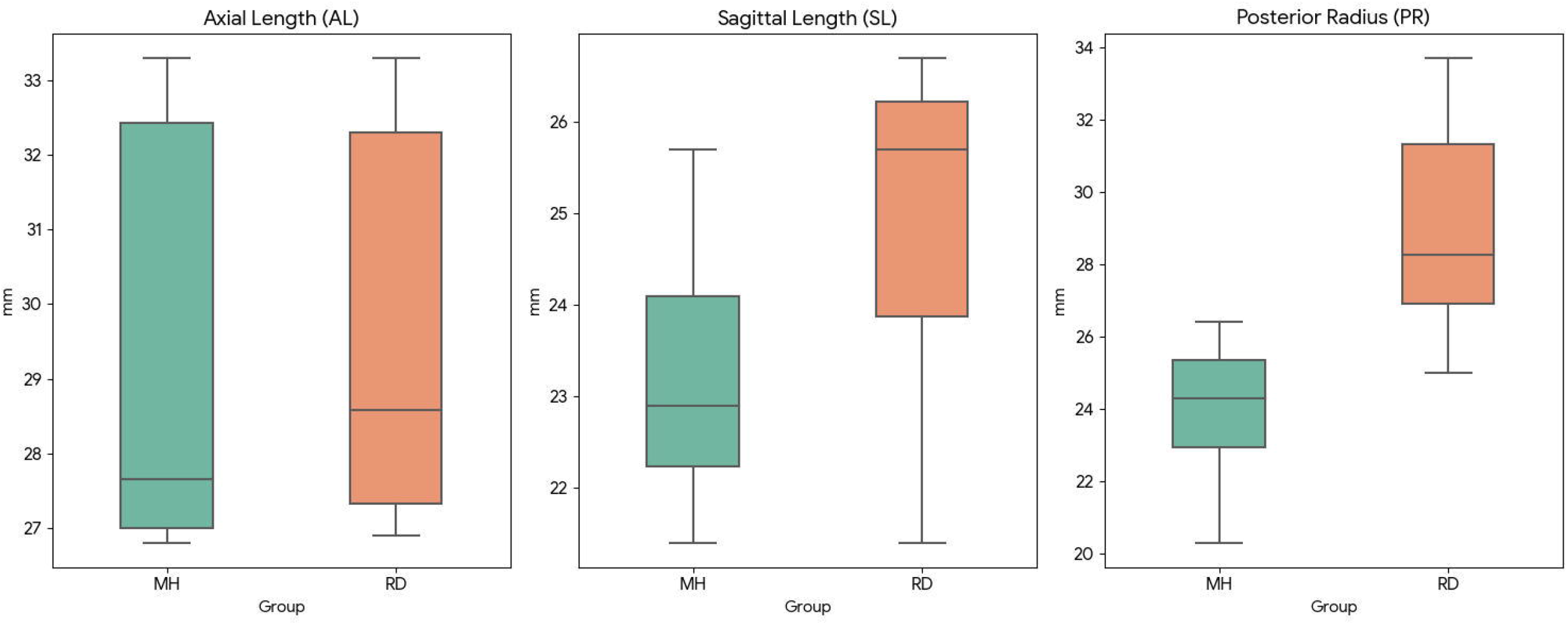
Illustration of the methods used to measure sagittal length (SL) and to calculate the radius of posterior curvature of the globe (rPC).

### Statistical Analysis

Normality was assessed using the Shapiro-Wilk test. Given the non-normal distribution of the samples, the Mann-Whitney U test was used to compare continuous variables. Fisher’s Exact Test was used to evaluate gender distribution. Data are presented as Median [Interquartile Range (IQR)]. Statistical significance was set at p<0.05.

## Results

The groups were statistically identical in age and sex. The MH group was 65% female, while the RD group was 45% female (p=0.341). Axial Length did not differ significantly between the groups (Median MH: 27.65 mm vs RD: 28.58 mm; p=0.39). Significant differences were found in the sagittal plane. The RD group showed a median SL of 25.70 mm, significantly larger than the 22.90 mm found in the MH group (p=0.001). Similarly, the rPC was significantly flatter in the RD group (Median: 28.25 mm) compared to the MH group (Median: 24.30 mm), with p < 0.001. This results are detailed in Table 1 and in Figure 2. A case comparison is shown in Figure 3.

**Table 1.**
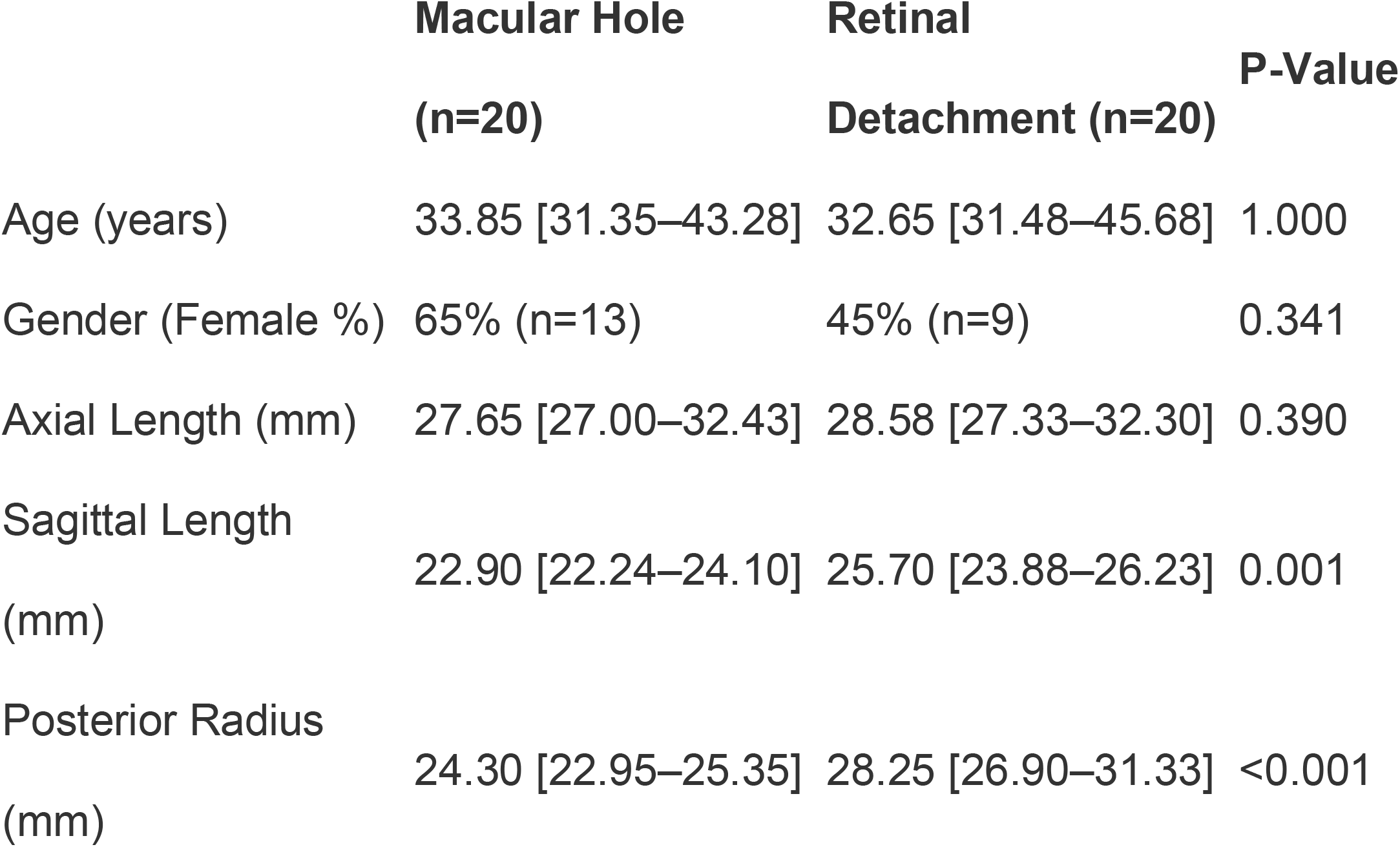
Study variables by group. Results are shown as median [interquartile range].

**Figure 2.**
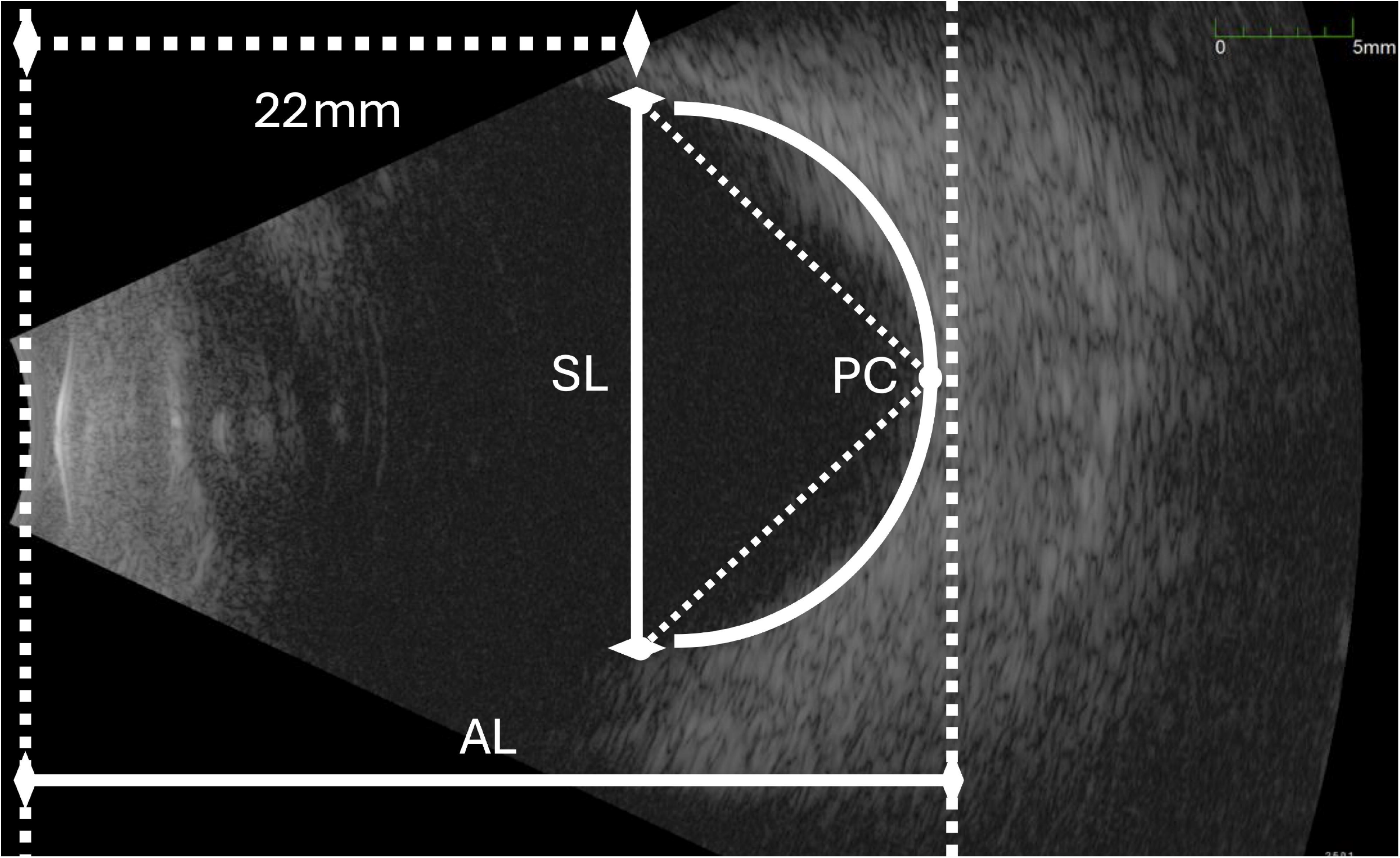
Box-plot showing the study variables by group.

**Figure 3.**
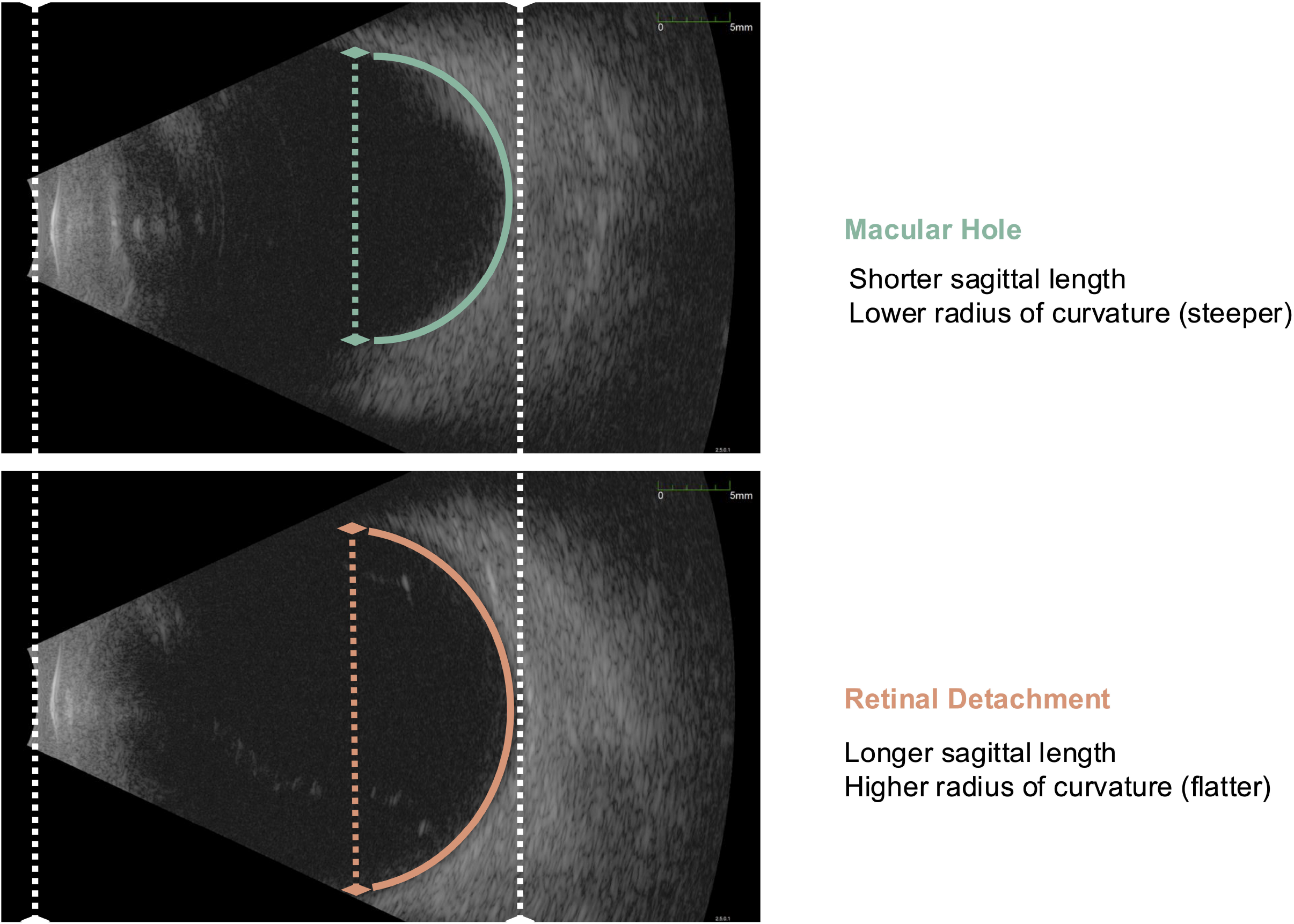
Case comparison between retinal detachment and macular hole eyes.

## Discussion

Our results demonstrate that eyes with retinal detachment possess a fundamentally different shape than those with macular holes, even when AL is the same. The flattening of the posterior curvature in the RD group (rPC = 28.25 mm) has profound biomechanical consequences. According to Laplace’s Law, wall tension is directly proportional to the radius of curvature [4]. Eyes in the RD group likely experience higher global wall tension, which, combined with the vertical expansion (high SL), increases mechanical stress on the peripheral retina [5]. This may explain the predisposition to peripheral tears in these patients. In contrast, the MH group has a more oval geometry (smaller PR and SL). In these eyes, pathologic forces are not distributed peripherally but are instead concentrated at the fovea, consistent with the anteroposterior vitreomacular traction described [6,7]. Furthermore, the globe shape has been shown to alter vitreous fluid dynamics during eye movement, potentially increasing shear stress in the RD group [8]. Furthermore, these shape variations are often associated with the development of posterior staphylomas in pathological myopia [9] and anomalous posterior vitreous detachment [10]. This study suggests that reliance on AL alone is insufficient for risk stratification. High SL and rPC flattening serve as markers the mechanics of retinal detachment. Dispite non.significant, there was a slighlty higer proportion of females in the MH group. This is noteworthy as some epidemiological studies suggest a higher prevalence of MH in women [3].

## Conclusion

The geometry of the globe in RD patients is characterized by significant vertical expansion and posterior flattening compared to MH patients. These findings emphasize the importance of three-dimensional biometric assessment in clinical practice.

## Data Availability

All data produced in the present study are available upon reasonable request to the authors

## References

1. Curtin, B. J. (1985). The myopias: Basic science and clinical management. Harper & Row.

2. Moriyama, M., Ohno-Matsui, K., Hayashi, K., Shimada, N., Yoshida, T., Tokoro, T., & Mochizuki, M. (2011). Topographic analyses of shape of eyes with pathologic myopia by high-resolution three-dimensional magnetic resonance imaging. Ophthalmology, 118(8), 1626–1637. 10.1016/j.ophtha.2011.01.018

3. Sigal, I. A., & Ethier, C. R. (2009). Biomechanics of the optic nerve head. Experimental Eye Research, 88(4), 799–807. 10.1016/j.exer.2009.02.003

4. Avitabile, T., Bonfiglio, V., Cicero, A., Torrisi, B., Reibaldi, M., Faro, S., & Longo, A. (2018). Correlation between axial length and retinal tears in eyes with posterior vitreous detachment. Retina, 38(2), 366–371. 10.1097/IAE.0000000000001540

5. Gaudric, A., Haouchine, B., Massin, P., Paques, M., Blain, P., & Erginay, A. (1999). Macular hole formation: New data on the pathogenesis based on optical coherence tomography. Archives of Ophthalmology, 117(6), 744–751. 10.1001/archopht.117.6.744

6. Spaide, R. F. (2020). Vitreous traction and macular holes. Retina, 40(3), 403– 405. 10.1097/IAE.0000000000002730

7. Meskauskas, J., Repetto, R., & Siggers, J. H. (2012). Shape change of the vitreous chamber during eye movements and its influence on vitreous motion. Investigative Ophthalmology & Visual Science, 53(10), 6123–6131. 10.1167/iovs.12-10041

8. Ohno-Matsui, K. (2014). Posterior staphyloma in pathologic myopia. Retina, 34(3), 453–463. 10.1097/IAE.0000000000000057

9. Sebag, J. (2004). Anomalous posterior vitreous detachment: A unifying concept in vitreo-retinal disease. Graefe’s Archive for Clinical and Experimental Ophthalmology, 242(8), 690–698. 10.1007/s00417-004-0980-y

10. Evans, J. R., Schwartz, S. D., McHugh, J. D., Casswell, A. G., Gregory-Evans, K., Goldberg, G. S., & Hykin, P. G. (1998). The epidemiology of idiopathic macular hole. Ophthalmology, 105(9), 1603–1606. 10.1016/S0161-6420(98)99026-9

